# Clinicogenomic analysis of breakthrough infections by SARS CoV2 variants after ChAdOx1 nCoV- 19 vaccination in healthcare workers

**DOI:** 10.1101/2021.06.28.21259546

**Authors:** Pratibha Kale, Ekta Gupta, Chhagan Bihari, Niharika Patel, Sheetalnath Rooge, Amit Pandey, Meenu Bajpai, Vikas Khillan, Partha Chattopadhyay, Priti Devi, Ranjeet Maurya, Neha Jha, Priyanka Mehta, Manish Kumar, Pooja Sharma, Sheeba Saifi, Aparna S, Sarfaraz Alam, Bharathram Uppili, Mohammed Faruq, Anurag Agrawal, Rajesh Pandey, Shiv Kumar Sarin

## Abstract

**Background:** India saw a massive surge and emergence of SARS CoV2 variants. We elucidated clinical and humoral immune response and genomic analysis of vaccine breakthrough (VBT) infections after ChAdOx1 nCoV-19 vaccine in healthcare workers (HCWs).

**Methods:** The study was conducted on 1858 HCWs receiving two doses of ChAdOx1 nCoV-19 vaccine. Serial blood samples were collected to measure SARS CoV2 IgG and neutralizing antibodies. 46 RT-PCR positive samples from VBT infections were subjected to whole genome sequencing (WGS).

**Results:** Infection was confirmed in 219 (11.79%) HCWs of which 21.46% (47/219) were non-vaccinated, significantly more (p <0.001) than 9.52% (156/1639) vaccinated group. VBT infections were significantly higher in doctors and nurses compared to other hospital staff (p <0.001). Unvaccinated individuals had 1.57 times higher risk of infection compared to partially vaccinated (p 0.02) and 2.49 times than fully vaccinated (<0.001). Partially vaccinated were at higher risk of infection than fully vaccinated (RR 1.58,p 0.01). There were 3 (1.36%) severe cases and 1 death in unvaccinated group compared to none in the vaccinated. Non-response after 14 days of second dose was seen in 6.5% (3/46) and low antibody levels (1-4.62 S/CO) in 8.69% (4/46). Delta variant (B.1.617.2) was dominant (69.5%) and reinfection was documented in 4 (0.06%) HCWs.

**Conclusions:** Nearly one in ten vaccinated HCWs can get infected, more so with only single dose (13.65%) than two doses (8.62%). Fully vaccinated are better protected with higher humoral immune response. Genomic analysis revealed an alarming rise of Delta variant (B.1.617.2) in VBT infections.

## Introduction

Vaccines have emerged as an effective countermeasure against the accelerating global expansion of severe acute respiratory syndrome coronavirus 2 (SARS-CoV-2) infection causing coronavirus disease 2019 (COVID-19). Multiple vaccine rollout across the world will curb the pandemic by protecting the vulnerable population groups.^1^ Vaccines are crucial for the individuals who fail to develop protective immunity, those with underlying medical or debilitating conditions, immunodeficiencies or elderly population. As the disease pathophysiology and effective immunity through vaccination is still an evolving science, we cannot predict protective immune response and breakthrough infections in vaccinated population.

In India till April 30^th^ 2021, the cumulative vaccinations administered were 15,48,54,096, including 94,10,892 in Healthcare Workers (HCWs) as 1st dose and 62,40,077 HCWs who had taken both the doses.^2^ There was a sudden surge in the COVID19 cases from mid-March to May 2021, with the worst affected states being Maharashtra and Delhi.^3^ This surge was predominantly caused by highly transmissible variants with potentials of immune evasion.^4^ Similar surge in cases was reported from the UK, South Africa and Brazil, and subsequently spreading world over. These SARS-CoV-2 variants of interest were namely Alpha (B.1.1.7; 501Y.V1), Beta (B.1.351; 501Y.V2) and Gamma; (B.1.1.28.1; 501Y.V3; P.1) respectively. India has reported the rapid spread of Alpha variant and witnessed the emergence of variant of concern Delta; (B.1.617.2) and variant of interest Kappa (B.1.617.1).^4^ Breakthrough (BT) infections after vaccination have been reported from USA at a rate of 0.01% and from a chronic care medical facility in India at a rate of 16.8%.^5,6^ In India, the ChAdOx1 nCoV-19/ AZD1222 (Covishield) vaccine was the main vaccine used across the country. We studied the VBT infections, their clinical characteristics, immune response and genomic analysis in a large cohort of vaccinated HCWs.

## Materials and methods

The study was conducted at the Institute of Liver and Biliary Sciences (ILBS), Delhi, India in collaboration with the CSIR-Institute of Genomics and Integrative Biology (CSIR-IGIB). The study had the approval of the Institutional Ethical Committee (IEC/2020/77/MA07) and was conducted following the Declaration of Helsinki. The study was conducted on 1858 HCWs at our university hospital from January to May 2021. Detailed demographic details and history of co-morbidities, prior history of COVID-19 infection, etc. were recorded.

For the purpose of this study the following definitions were followed:

1. *Vaccine Breakthrough infection (VBT):* A vaccine breakthrough infection was defined as the detection of SARS-CoV-2 RNA or antigen in a respiratory specimen collected from an individual who had received either one or two doses of vaccine.
2. *Partially vaccinated HCW:* The HCW who had received a single dose of vaccine or who developed infection before 14 days of the second dose was considered partially vaccinated.
3. *Fully vaccinated HCW:* The HCW who received two doses of vaccine and developed infection after 14 days of the second dose.
4. *Reinfection:* Reinfection was defined as detection of SARS-CoV-2 RNA on or after 90 days of the first detection of SARS-CoV-2 RNA and paired respiratory specimens (one from each infection episode) were available.
5. *Non-responder:* HCW with no detectable SARS COV2 IgG antibody response 14 days after the second dose of the vaccine.
6. *Low antibody levels:* SARS COV2 IgG antibodies detectable in the range of 1 to 4.62 signal/cut-off (S/CO) were considered Low level antibody response.
7. *Medium antibody levels:* SARS COV2 IgG antibodies detectable in the range of more than 4.62 to 18.45 S/CO were considered medium level antibody response.
8. *High antibody levels:* SARS COV2 IgG antibodies detectable in the range of >18.45 S/CO were considered high level antibody response.

### 1. Humoral immune response

Blood samples were collected at baseline before vaccination and then 28 days following the first dose, at second dose and after 14 days. The SARS COV2 IgG antibodies were measured using the enhanced chemiluminescence method (Vitros ECi, Ortho Clinical Diagnostics, New Jersey, US). This a qualitative assay based on a recombinant form of the SARS-CoV-2 spike subunit 1 protein. Results are based on the sample signal-to-cut-off (S/Co) ratio, with values <1.0 as negative and ≥1.00 as positive results. Further the presence of neutralizing antibodies were measured by surrogate neutralization ELISA. (Genscript, USA).

### 2. Confirmation of breakthrough infection

SARS CoV2 infection was confirmed in 203 HCWs during the study period. Combined nasopharyngeal and oral swabs were collected in viral transport media (VTM) from all the symptomatic study subjects. Samples were tested for the presence of dual genes E and RdRP by real time reverse transcriptase polymerase chain reaction(RT-PCR) using commercial kit (Q-Line®, M/s POCT Pvt Ltd.) Samples with detection of both the genes with < 35 cycle threshold (Ct) value were considered as positive.

### 3. SARS-CoV-2 Whole genome sequencing

#### Sequencing using Illumina MiSeq Platform

46 RT-PCR positive samples (Ct value ≤ 20) were randomly selected from the Virology repository at -80°C and subjected to whole genome sequencing (WGS). WGS of the RNA elutes was carried out using the Genome Analyzer IIx (Illumina, San Diego, USA) according to the manufacturer’s instructions. Double-stranded cDNA (ds cDNA) was synthesised from the RNA elutes. The first strand of cDNA was synthesised using Superscript IV First strand synthesis system (Thermo Fisher Scientific, Cat.No. 18091050) followed by single-stranded RNA (ssRNA) digestion with RNase H for second strand synthesis using DNA Polymerase I Large (Klenow) Fragment (New England Biolabs, Cat. No. M0210S). The ds cDNA was purified using AMPure XP beads (AMPure XP, Beckman Coulter, Cat. No. A63881) and quantified using NanoDrop (ND-1000 UV-Vis Spectrophotometer, Thermo Fisher Scientific). 100ng of purified ds cDNA was used for library prep using the Illumina DNA Prep with Enrichment kit (Illumina, Cat. No. 20018705). The process involves tagmentation followed by cleanup and amplification leading to indexed DNA fragments. Following tagmentation and indexing, enrichment was performed using the Illumina RVOP (Illumina, Cat no. 20042472), wherein 500ng of each sample were pooled by mass in accordance with the reference guide (Illumina, Doc. No. 1000000048041v05) for the 12-plex hybridisation with biotinylated adjacent oligo-probes of the RVOP. The hybridisation was performed overnight after which the probes were captured by streptavidin-biotin based interactions. The final library was PCR amplified and purified before sequencing. The quality and quantity of the sequencing library was checked using Agilent 2100 Bioanalyzer with high sensitivity DNA chip (Catalog number: 5067-4626) and the Qubit dsDNA HS Assay kit (Catalog number: Q32851), respectively. A loading concentration of 10pM was prepared by denaturing and diluting the libraries in accordance with the MiSeq System Denature and Dilute Libraries Guide (Illumina, Document no. 15039740 v10). Sequencing was performed on the MiSeq system, using the MiSeq Reagent Kit v3 (150 cycles) at 2 x 75 bps read length.

### 4. Sequencing data analysis

#### MiSeq data analysis

FastQC v0.11.9 (http://www.bioinformatics.babraham.ac.uk/projects/fastqc) was used to check the Phred quality score for all sequences. For all samples, the quality score threshold was 20 and above. Adapter trimming was performed using the Trim Galore tool v0.6.1 (https://www.bioinformatics.babraham.ac.uk/projects/trim_galore/) and alignment of sequences was performed using the HISAT2 [7] algorithm on human data build GRCh38. To remove any human sequences from the dataset, samtools v1.12 [8] were used to remove aligned sequences. Henceforth, only unaligned sequences were taken into consideration. BCFTools v1.12 was used to generate consensus fasta and variant calling.

### 5. SARS-CoV-2 Phylogenetic and mutation analysis

We sequenced 46 vaccinated COVID-19 positive samples that were aligned to NC_045512 reference genome using MAFFT v7.475 [9] multiple alignment tool. The aligned sequences were trimmed to remove gaps and a phylogenetic tree was generated using the default model of the IQ-TREE tool v2.0.3 [10]. The tree was visualized using FigTree v1.4.4 [11]. Further, the assembled SARS-CoV-2 genomes were assigned lineages using the package Phylogenetic Assignment of Named Global Outbreak LINeages (PANGOLIN) [12]. The lollipop plot is generated in RStudio using g3viz, rtracklayer, and trackViewer packages followed by data visualization using the ggplot2 package. All the figures were updated using Inkscape software [13].

#### Data Availability

The consensus fasta generated for this study has been submitted in GISAID under the accessions: EPI_ISL_2424135 = 1, EPI_ISL_2426145 to EPI_ISL_2426189 = 45

#### Data analysis

The collected data was entered in excel spreadsheet and expressed as median or as percentage. The categorical data were analysed using Chi-Square or Fisher’s exact test. The statistical analysis was done using SPSS software version 22.

## Results

### Baseline characteristics of infections in HCWs

A total of 1858 HCWs were enrolled in the study. The HCWs were divided in two groups vaccinated (88.2%, 1639/1858) and non-vaccinated (11.7%, 210/1858). (Table1) The vaccinated group was further subdivided as partially vaccinated (17.8%, 293/1639) and fully vaccinated (82.12%, 1346/1639). Overall SARS CoV2 infections was seen in 219 (11.79%) HCWs during the study period. 21.46% (47/219) infections were documented in non-vaccination group, significantly more (p <0.001) than the vaccinated group (Table 1).. Breakthrough (BT) infections were confirmed in 9.52% (156/1639) of the vaccinated group. BT in partially vaccinated were significantly higher than in fully vaccinated HCWs; 13.65% (40/293) versus 8.62% (116/1346) (p= 0.008).

**Table 1.** Distribution of BT infections as per work profile of HCW:

| Category of HCW | Vaccinated<br>N= 1639 | Partially vaccinated<br>N= 293 | Fully vaccinated<br>N= 1346 | Non-vaccinated<br>N= 219 | TOTAL<br>N= 1858 | Level of significance<br>p value |
| --- | --- | --- | --- | --- | --- | --- |
| Doctors<br>n/N (%) | 30/129<br>(23.26) | 8/29<br>(27.59) | 22/100<br>(22) | 5/18<br>(27.78) | 35/147<br>(23.81) | 0.754 |
| Nursing staff<br>n/N (%) | 76/314<br>(24.20) | 21/87<br>(24.14) | 55/227<br>(24.23) | 27/110<br>(24.55) | 103/424<br>(24.29) | 0.997 |
| Technicians<br>n/N (%) | 16/215<br>(7.44) | 5/60<br>(8.33) | 11/155<br>(7.10) | 4/20<br>(20) | 20/235<br>(8.51) | 0.150 |
| Non-Medical<br>n/N (%) | 14/321<br>(4.36) | 3/27<br>(11.11) | 11/294<br>(3.74) | 6/36<br>(16.67) | 20/357<br>(5.60) | **0.003\*** |
| GDA<br>n/N (%) | 20/660<br>(3.03) | 3/90<br>(3.33) | 17/570<br>(2.98) | 5/35<br>(14.29) | 25/695<br>(3.60) | **0.002\*** |
| Total<br>n/N (%) | 156/1639<br>(9.52) | 40/293<br>(13.65) | 116/1346<br>(8.62) | 47/219<br>(21.46) | 203/1858<br>(10.93) | **non-Vaccinated Vs vaccinated <0.001 Partially vaccinated** |
|  |  |  |  |  |  | **Vs Fully vaccinated**<br>**p = 0.008** |
| **p value** |  | **<0.001<sup>#</sup>** | **<0.001<sup>#</sup>** | 0.621 | **<0.001<sup>#</sup>** |  |

N = total number subjects in the category, n= number of subjects with infection in the category,
GDA: General duty assistant and housekeeping staff
\* = fully vaccinated Vs non-vaccinated and partially vaccinated Vs non-vaccinated have significant difference. No significant difference between fully vaccinated and partially vaccinated subjects

#### Distribution of BT infections as per work profile of HCW

Based on the work profile the distribution of infections in non-vaccinated staff was doctors 27.7% (5/18), nurses 24.5% (27/110), technicians 20% (4/20), non-medical staff 16.6% (6/36) and 14.28% (5/35) in general duty assistants (GDA). (Table 1) (Figure 1). BT infections in partially vaccinated individuals were 13.65% (40/293); doctors 27.59%, nurses 24.14%, technicians 8.33%, non-medical staff 11.11%, and 3.33% in GDA. Doctors and nurses had significantly higher number of infections as compared to technical staff, non-medical staff and GDA in the partially vaccinated group (p<0.001). BT infections documented in fully vaccinated group; 22% (22/100) in doctors, 24.2% (55/227) in nursing staff, 7.09% (11/155) in technical staff, 3.74% (11/294) in non-medical staff and 1.22% (17/553) in GDA. (Table 1) (Figure1). Doctors and nurses in the fully vaccinated group were significantly more prone to acquire BT as compared to technical staff, non-medical staff and GDA (P<0.001).

**Figure 1:**
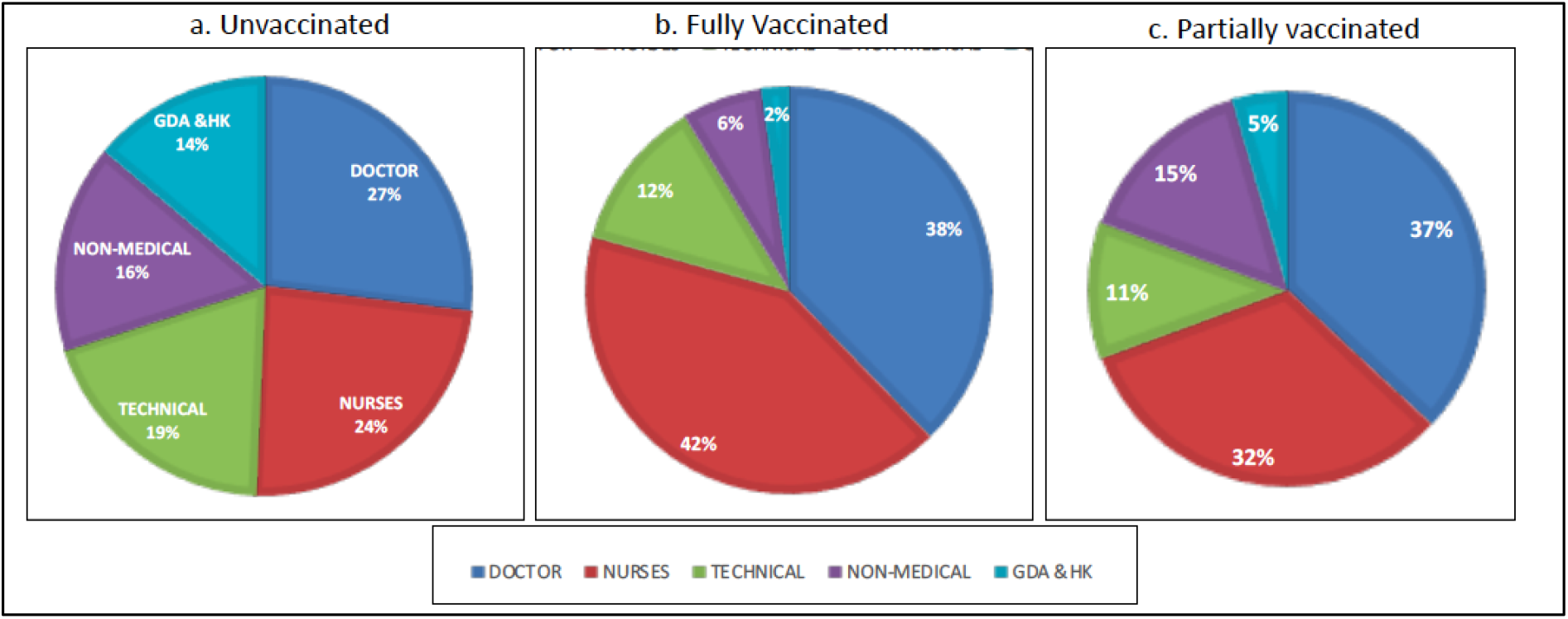
Distribution of SARS CoV2 infection in HCWs as per work profile, expressed as percentage. a. non-vaccinated HCWs, b. Fully vaccinated HCWs, c. partially vaccinated HCWs.

### Reinfection

Reinfection was documented in 4 HCWs, 6-8 months after the previous infection in the vaccinated group. These included doctors (2), who had received 2 doses of vaccine and nursing staff (1) and non-medical staff (1) who had received single dose of vaccine. The viral load was low in first 3 (Ct value 33.10, 29.92, 29.52) and high in the remainder (Ct value 20.52). All had mild symptoms of fever and body ache and no significant lung involvement.

#### Clinical presentation of BT infections

The median age was 34 (IQR: 21-67) years, with similar gender distribution, male 44.8% (91/203) and female 55.17% (112/203). Time to occurrence of BT infection in the recipients of both doses was 50 (IQR: 2-90) days and 30.5 (IQR: 3-95) days after single dose. Majority of infections were clinically mild with fever, bodyache, cough, headache, diarrhoea or vomiting with respiratory rate < 24/min and SpO2: ≥ 94% on room air. The non-vaccinated subjects were at a significantly higher risk of developing infection as compared to partial (RR 1.57, (95% CI 1.07-2.31) p=0.02) and fully vaccinated subjects (RR 2.49 (95%CI 1.83-3.39) p=< 0.001). Partially vaccinated subjects were at a higher risk of developing infection compared to fully vaccinated group (RR 1.58 (95% CI, 1.13-2.22) p=0.01). The risk of developing mild infection was equal in the non-vaccinated and partially vaccinated group (RR 1.0 (95% CI 0.6-1.65). However, both these groups had higher risk than the fully vaccinated group (RR 1.30 (95% CI 0.86-1.98), 1.30 (0.90-1.89 respectively). Similarly, the risk of developing moderate infection was also significantly higher in the unvaccinated than vaccinated subjects (RR 3.35 (95% CI 1.5-7.45) p=0.002.). None of the vaccinated subjects had severe infection requiring ICU admission and no death was reported. There were 3 (1.36%) severe cases and 1 death in the unvaccinated group. There was no significant association seen in co-morbidities with the BT.

#### Clinicogenomic association of BT infections

Genomic sequencing and analysis were available in 46 samples. We analysed the humoral immune response post vaccination in them (Table 3). Thirty-two (78.26%) had received 2 doses of vaccine and 14 (30.4%) received single dose.

**Table 2.**
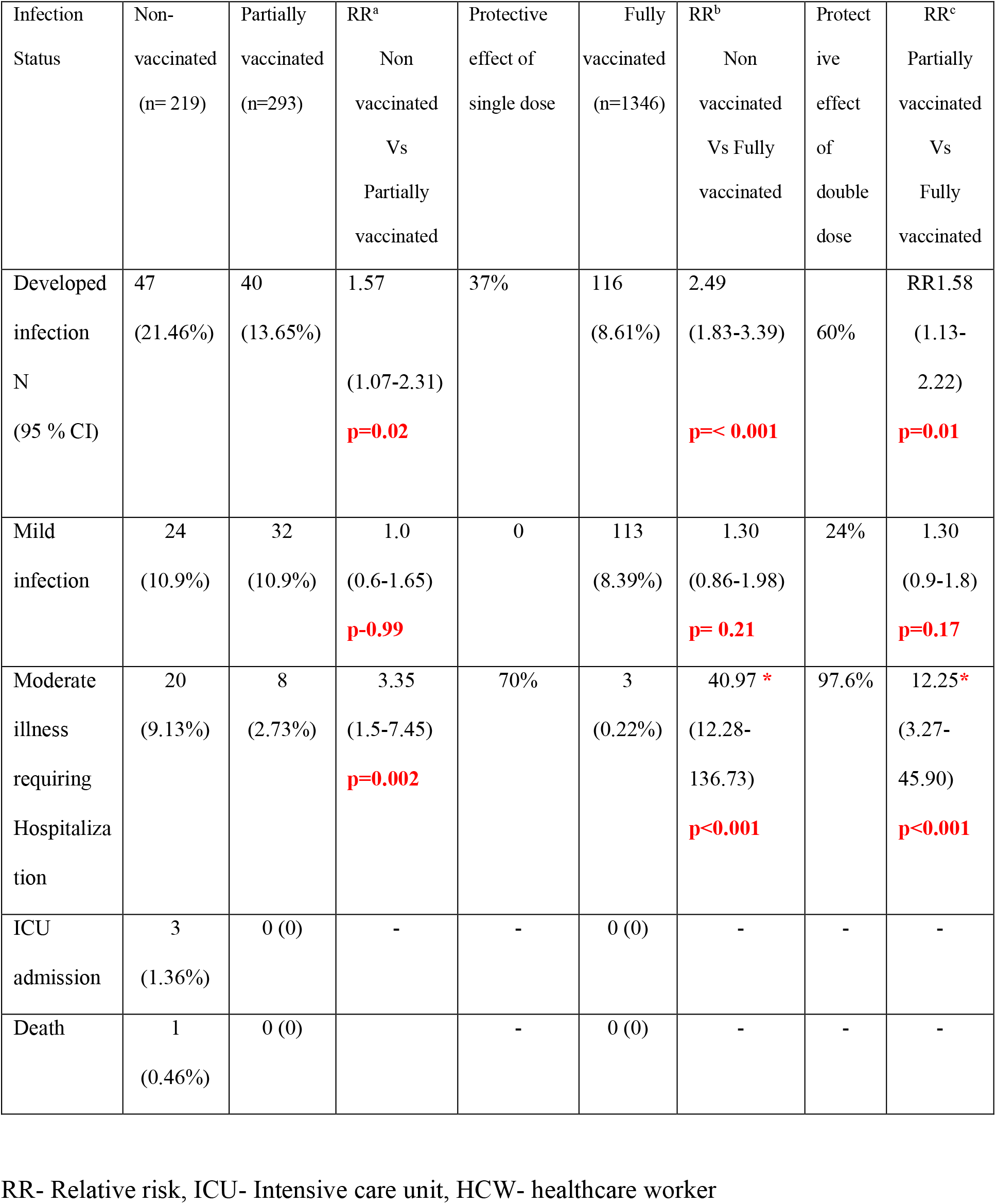
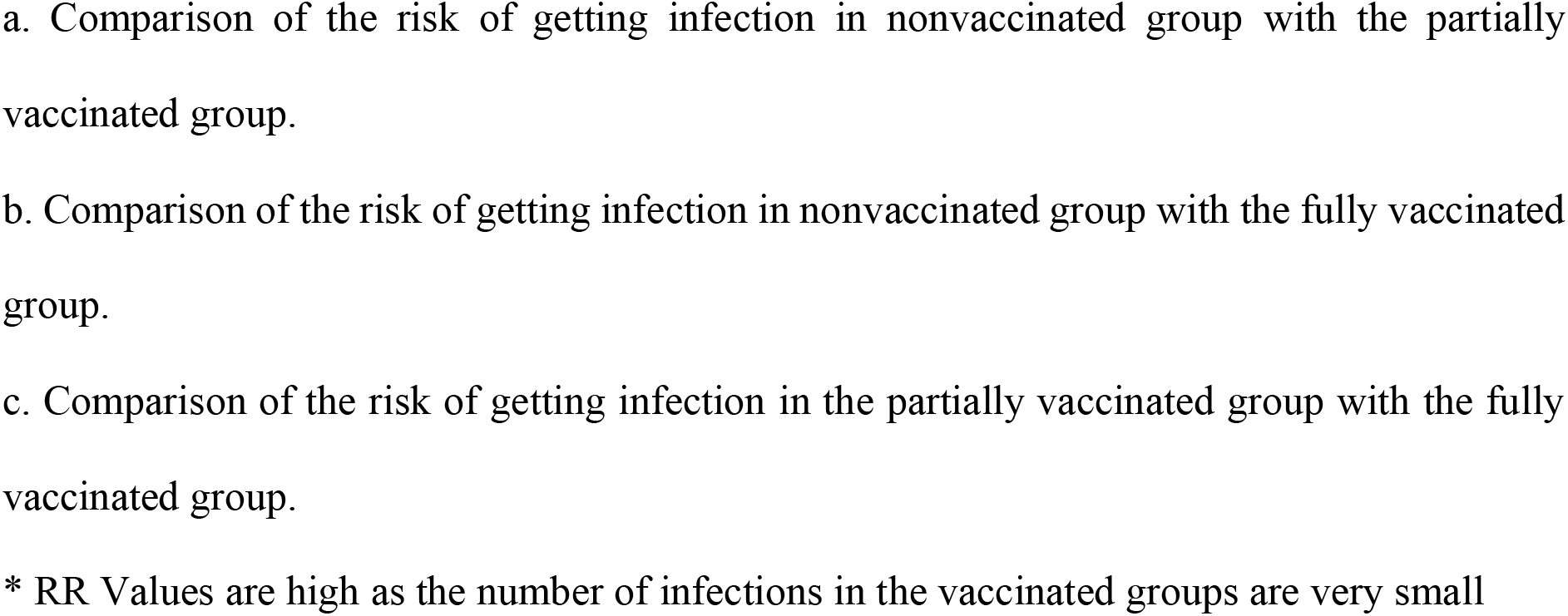
Clinical presentation of BT infections in HCWs and association with vaccination.

| Infection Status | Non-vaccinated<br>(n= 219) | Partially vaccinated<br>(n=293) | RR<sup>a</sup><br>Non vaccinated<br>Vs<br>Partially vaccinated | Protective effect of single dose | Fully vaccinated<br>(n=1346) | RR<sup>b</sup><br>Non vaccinated<br>Vs Fully vaccinated | Protective effect of double dose | RR<sup>c</sup><br>Partially vaccinated<br>Vs<br>Fully vaccinated |
| --- | --- | --- | --- | --- | --- | --- | --- | --- |
| Developed infection<br>N<br>(95 % CI) | 47<br>(21.46%) | 40<br>(13.65%) | 1.57<br>(1.07-2.31)<br>**p=0.02** | 37% | 116<br>(8.61%) | 2.49<br>(1.83-3.39)<br>**p=< 0.001** | 60% | RR1.58<br>(1.13-2.22)<br>**p=0.01** |
| Mild infection | 24<br>(10.9%) | 32<br>(10.9%) | 1.0<br>(0.6-1.65)<br>**p=0.99** | 0 | 113<br>(8.39%) | 1.30<br>(0.86-1.98)<br>**p= 0.21** | 24% | 1.30<br>(0.9-1.8)<br>**p=0.17** |
| Moderate illness requiring Hospitalization | 20<br>(9.13%) | 8<br>(2.73%) | 3.35<br>(1.5-7.45)<br>**p=0.002** | 70% | 3<br>(0.22%) | 40.97 *<br>(12.28-136.73)<br>**p<0.001** | 97.6% | 12.25*<br>(3.27-45.90)<br>**p<0.001** |
| ICU admission | 3<br>(1.36%) | 0 (0) | - | - | 0 (0) | - | - | - |
| Death | 1<br>(0.46%) | 0 (0) |  | - | 0 (0) | - | - | - |

RR- Relative risk, ICU- Intensive care unit, HCW- healthcare worker
- a. Comparison of the risk of getting infection in nonvaccinated group with the partially vaccinated group. - b. Comparison of the risk of getting infection in nonvaccinated group with the fully vaccinated group. - c. Comparison of the risk of getting infection in the partially vaccinated group with the fully vaccinated group.
\* RR Values are high as the number of infections in the vaccinated groups are very small

**Table 3:**
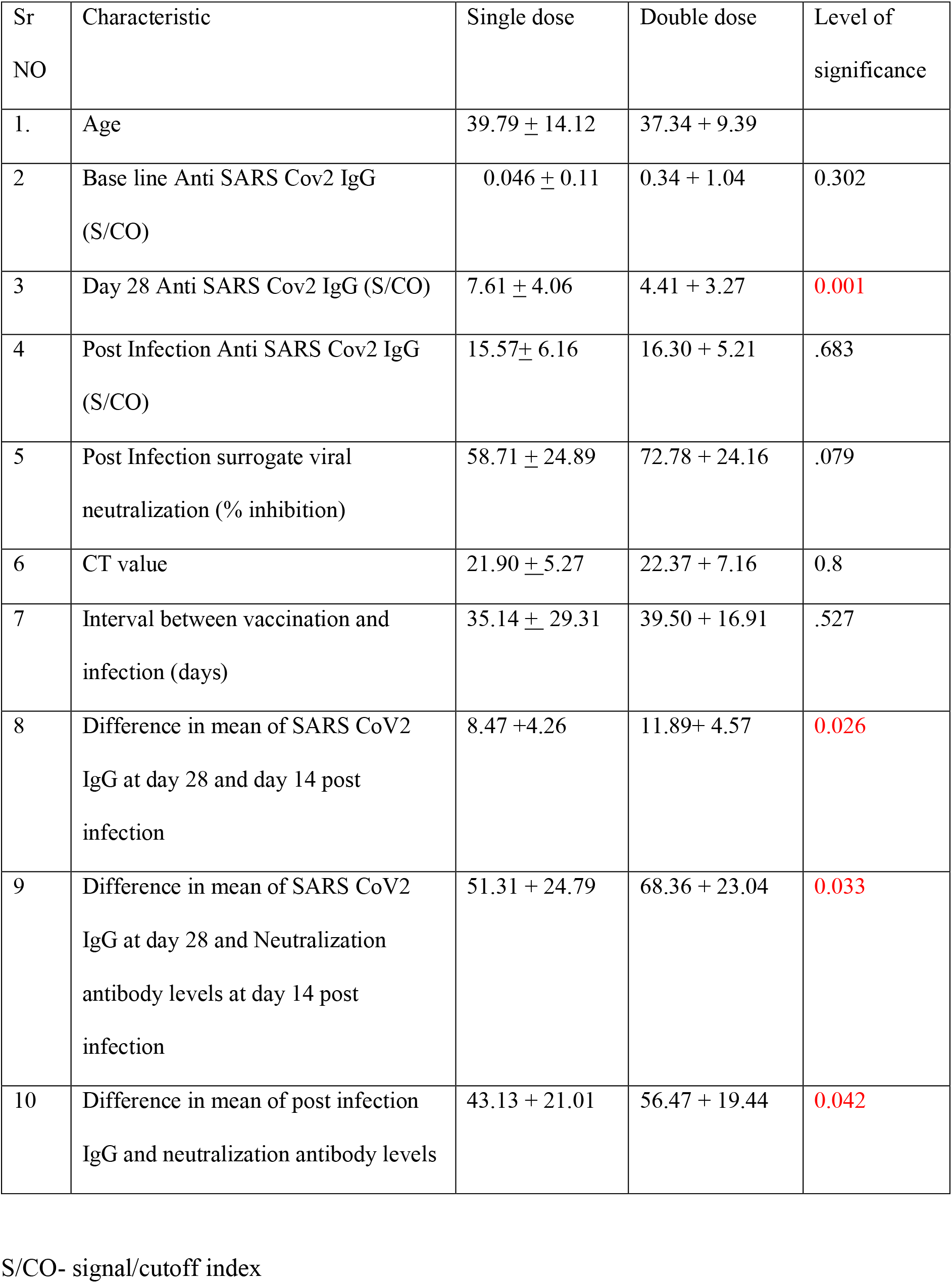
Characteristics of the 46 breakthrough infections.

#### Viral load

The approximate burden of viral load as measured indirectly by Ct value was almost similar in both the groups and did not differ with the vaccination status. The median Ct value was 21.13 (IQR 11.95-29.52) in partially vaccinated and 23.24 (IQR 0.00-33.10) in vaccinated group (p value : 0.827)

#### IgG antibody levels at baseline and day 14

Baseline IgG levels were checked in the study group in order to understand any prior infection and its effect on the attainment of post vaccination immune response.

The IgG antibody levels at baseline (before vaccination) were negative (<1 signal/cut-off, S/CO) in 93.4% (43/46), 3 HCWs had low level IgG antibodies (3.04-4.32 S/CO) at baseline. There was no difference in the baseline IgG antibodies between the partially and fully vaccinated HCWs (0.046 ± 0.11 vs 0.34 ± 1.04, p = 0.302 (S/CO, mean ± SD). The antibody levels at day 14 were also comparable in both the groups (0. 436 ± 0. 756 vs 1.497 ± 2.492, p= 0.128). (Table 1) (Figure 2)

**Fig 2:**
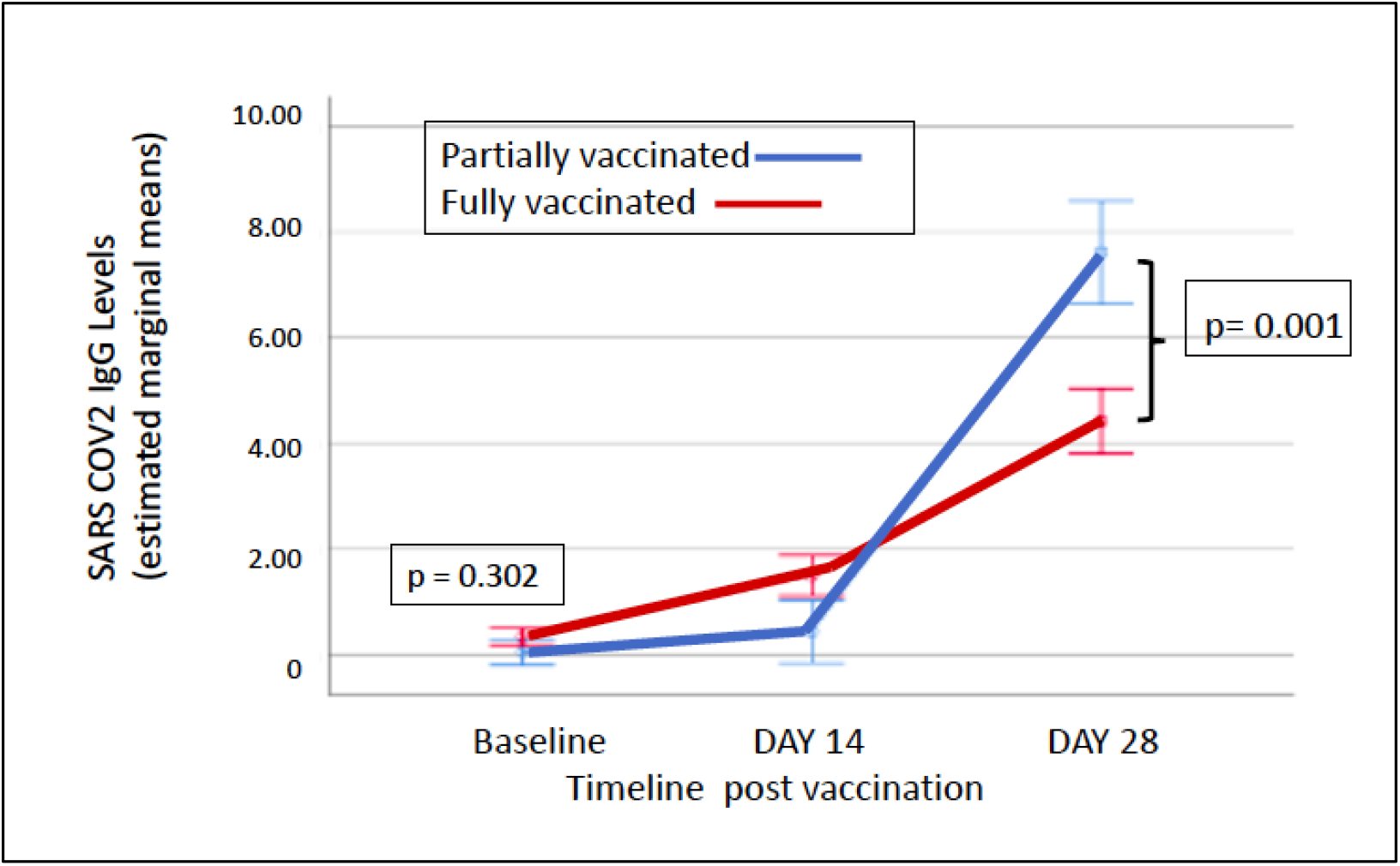
Difference in the SARS CoV2 IgG antibody levels at 28 days in partially and fully vaccinated HCWs.

#### 28 day IgG antibody level

During the initial period of vaccination drive from January to March, the recommended gap between two doses was 4-6 weeks. Hence the study protocol included sample collection at 28 days. At this time point, rise in antibodies was significantly higher in partially vaccinated group (7.61 ± 4.06 vs 4.41 ± 3.27, p= 0.001 (S/CO, mean ± SD) (Fig 2).

#### Antibody response after second dose

The antibody levels were tested 14 days after the second dose and showed; non-response in 6.5% (3/46), low antibody levels (1-4.62 S/CO) in 8.69% (4/46), medium (4.62-18.45 S/CO) in 50% (23/46). No subject had high (>18.45 S/CO) levels of antibodies.^6^ There was a significant difference in the IgG antibody and neutralizing antibody levels at 14 days post infection in the 2 groups (table 3). The levels were significantly higher in recipients of the second dose for difference in the mean change from day 28 IgG after vaccination to post-vaccination levels of IgG and the neutralizing antibodies (8.47 ±4.26 Vs 11.89± 4.57, p= 0.026, 51.31 ± 24.79vs 68.36 ± 23.04,p=0.033) respectively. (Table 3, fig 3)

**Figure3:**
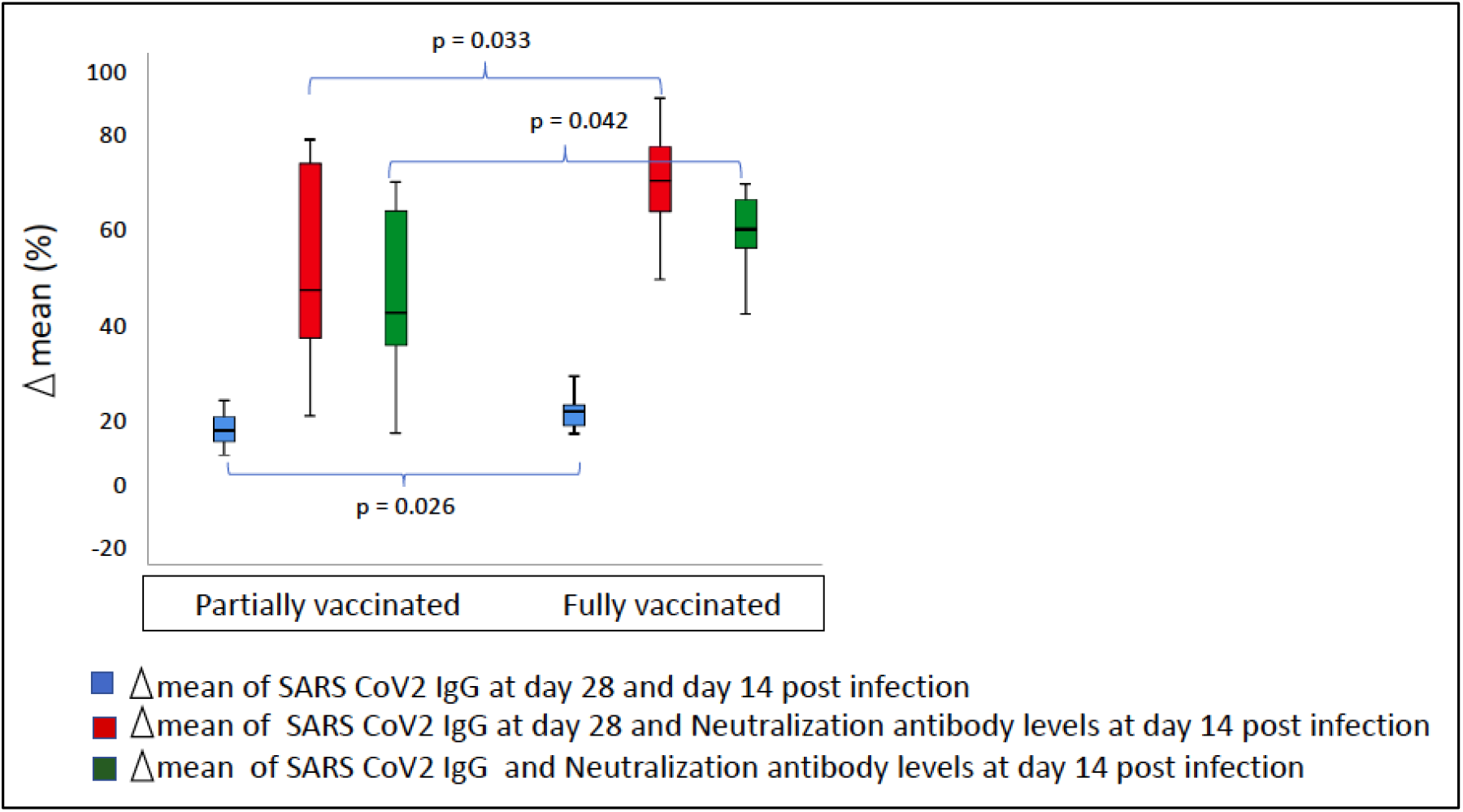
Box and whisker plot to show the difference in mean (Δ mean) in IgG antibody levels day 28 and day 14 post infection, IgG antibody levels day 28 and neutralization day 14 post infection and IgG antibody levels and Neutralizing antibodies day 14 post infection.

#### Phylogenetic and mutation analysis

The presence of the B.1.617 lineage was found in abundance in a phylogenetic examination of 46 vaccinated COVID-19 positive samples. (Figure 4).The B.1.617.2 (delta variant) (n=32; 69.56%), B.1.617.1 (kappa variant) (n=11; 23.91 %), and only one patient with B.1.1.7 strain (alpha variant) were identified (Figure 5).

**Figure 4:**
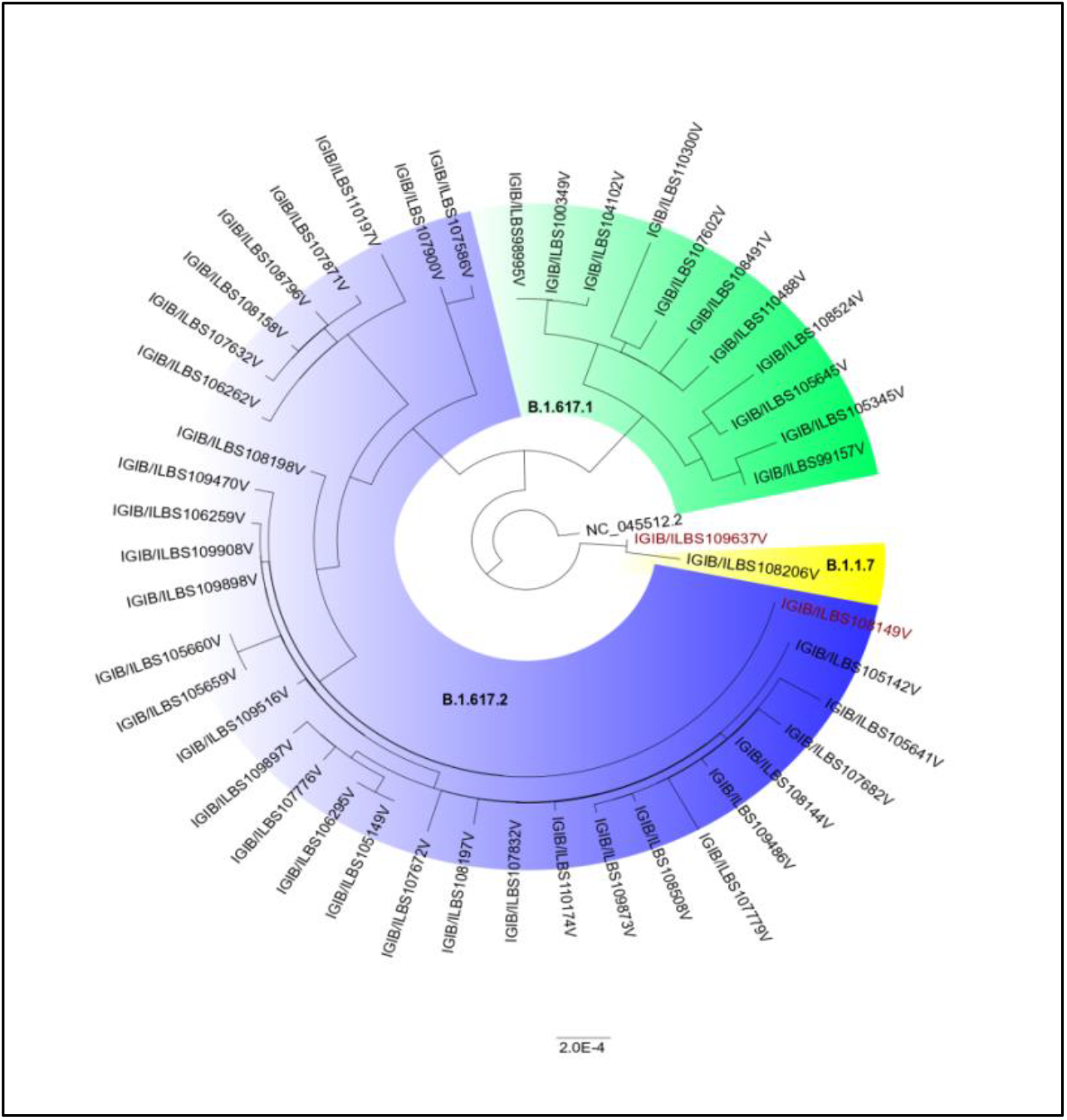
The Phylogenetic distribution of lineages in 46 vaccinated samples

**Figure 5:**
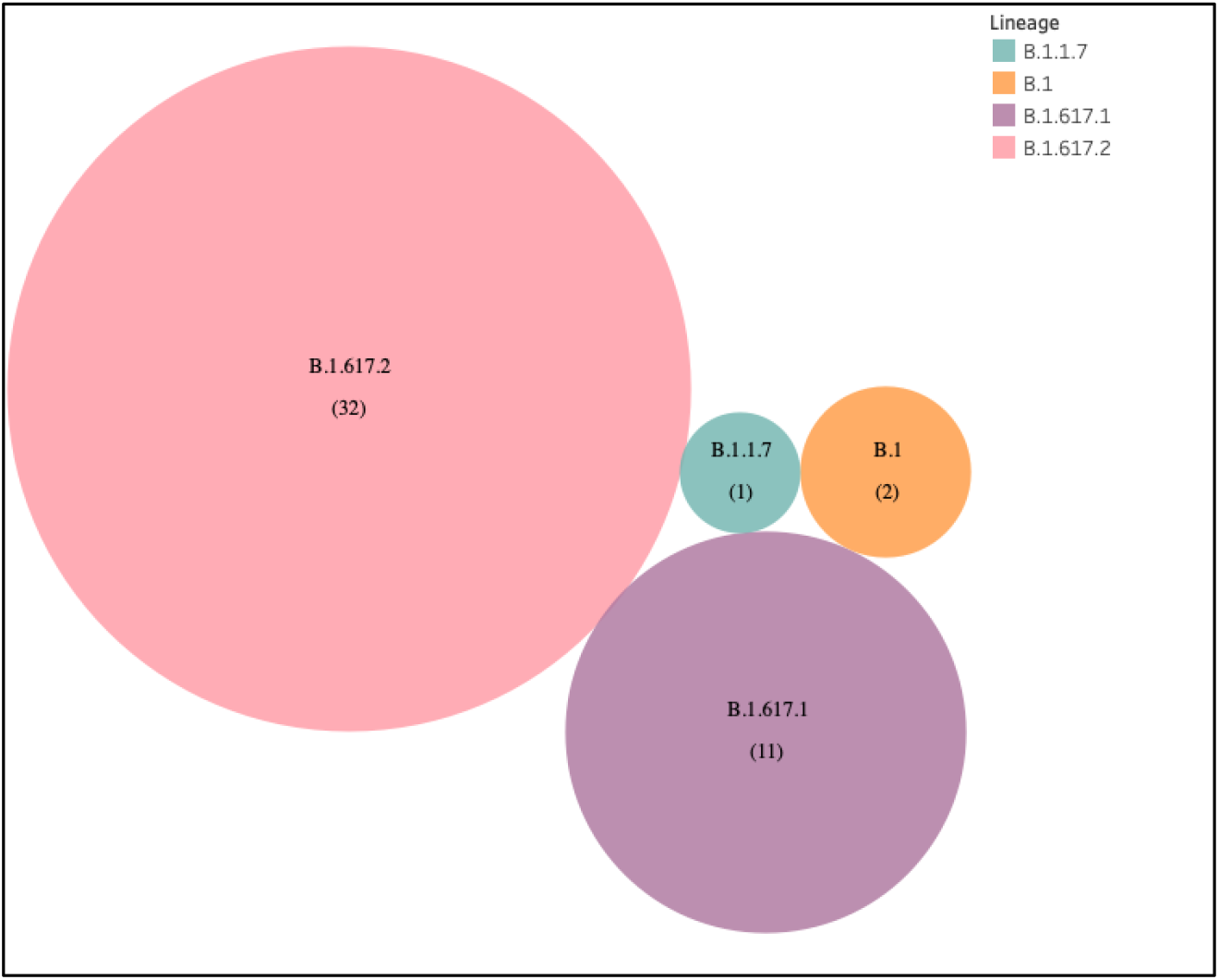
Prevalence of B.1.617.2 variants in post-vaccinated samples.

The presently widespread B.1.617.2 lineage-defining mutations were detected in significant frequency in our samples when we looked at the top most frequent mutations. C23604G (S: P681R) mutation occurred in 95.65 percent of all sequences, whereas T22917G (S: L452R) and C22995A (S: T478K) mutations occurred in 93.48 percent and 63.04 percent, respectively. (Figure 6). Other commonly occurring mutations include A23403G (S: D614G), G28881T (N:203), and G29402T (N:377). While the majority of these changes were non-synonymous, one synonymous mutation, C3073T, was found in all of the samples (Table 4). The Kappa variant was significantly associated with moderate cases in partially vaccinated HCWs (p=0.04) (Table 5)

**Figure 6:**
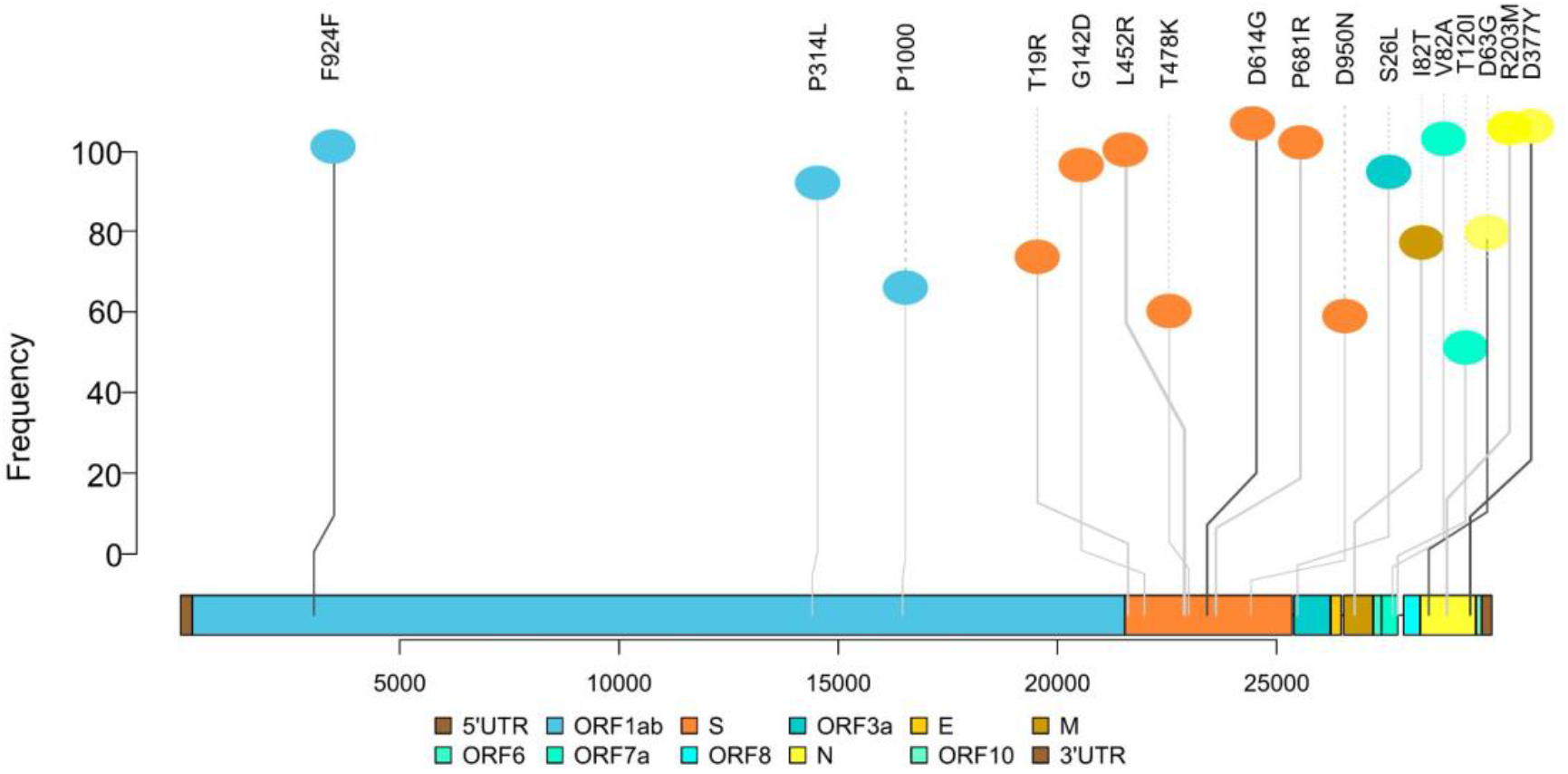
Top most frequent mutations detected in 46 vaccine-breakthrough samples.

**Table 4:** Top frequently occurring mutations in the study

| Position | Variants | AA_mutation | Gene | Count | Percentage | Type of Mutations |
| --- | --- | --- | --- | --- | --- | --- |
| 3037 | C3037T | F924F | ORF1a:924 | 46 | 100.00 | synonymous |
| 23403 | A23403G | D614G | S:614 | 46 | 100.00 | non-synonymous |
| 23604 | C23604G | P681R | S:681 | 44 | 95.65 | non-synonymous |
| 22917 | T22917G | L452R | S:452 | 43 | 93.48 | non-synonymous |
| 28881 | G28881T | R203M | N:203 | 43 | 93.48 | non-synonymous |
| 29402 | G29402T | D377Y | N:377 | 43 | 93.48 | non-synonymous |
| 27638 | T27638C | V82A | ORF7a | 41 | 89.13 | non-synonymous |
| 14408 | C14408T | P314L | ORF1b:314 | 40 | 86.96 | non-synonymous |
| 25469 | C25469T | S26L | ORF3a:26 | 40 | 86.96 | non-synonymous |
| 21987 | G21987A | G142D | S:142 | 38 | 82.61 | non-synonymous |
| 21618 | C21618G | T19R | S:19 | 31 | 67.39 | non-synonymous |
| 16466 | C16466T | P1000L | ORF1b:1000 | 30 | 65.22 | non-synonymous |
| 26767 | T26767C | I82T | M:82 | 30 | 65.22 | non-synonymous |
| 28461 | A28461G | D63G | N:63 | 30 | 65.22 | non-synonymous |
| 22995 | C22995A | T478K | S:478 | 29 | 63.04 | non-synonymous |
| 24410 | G24410A | D950N | S:950 | 27 | 58.70 | non-synonymous |
| 27752 | C27752T | T120I | ORF7a:120 | 27 | 58.70 | non-synonymous |

#### Overlap of infections in HCWs with the community surge in cases

The COVID-19 cases reported in Delhi during the study period were retrieved. The clustering of cases in the HCWs overlapped with the surge in the cases in the community. The number of cases, viral load and the antibody response in these cases is depicted in figure 7.

**Fig 7:**
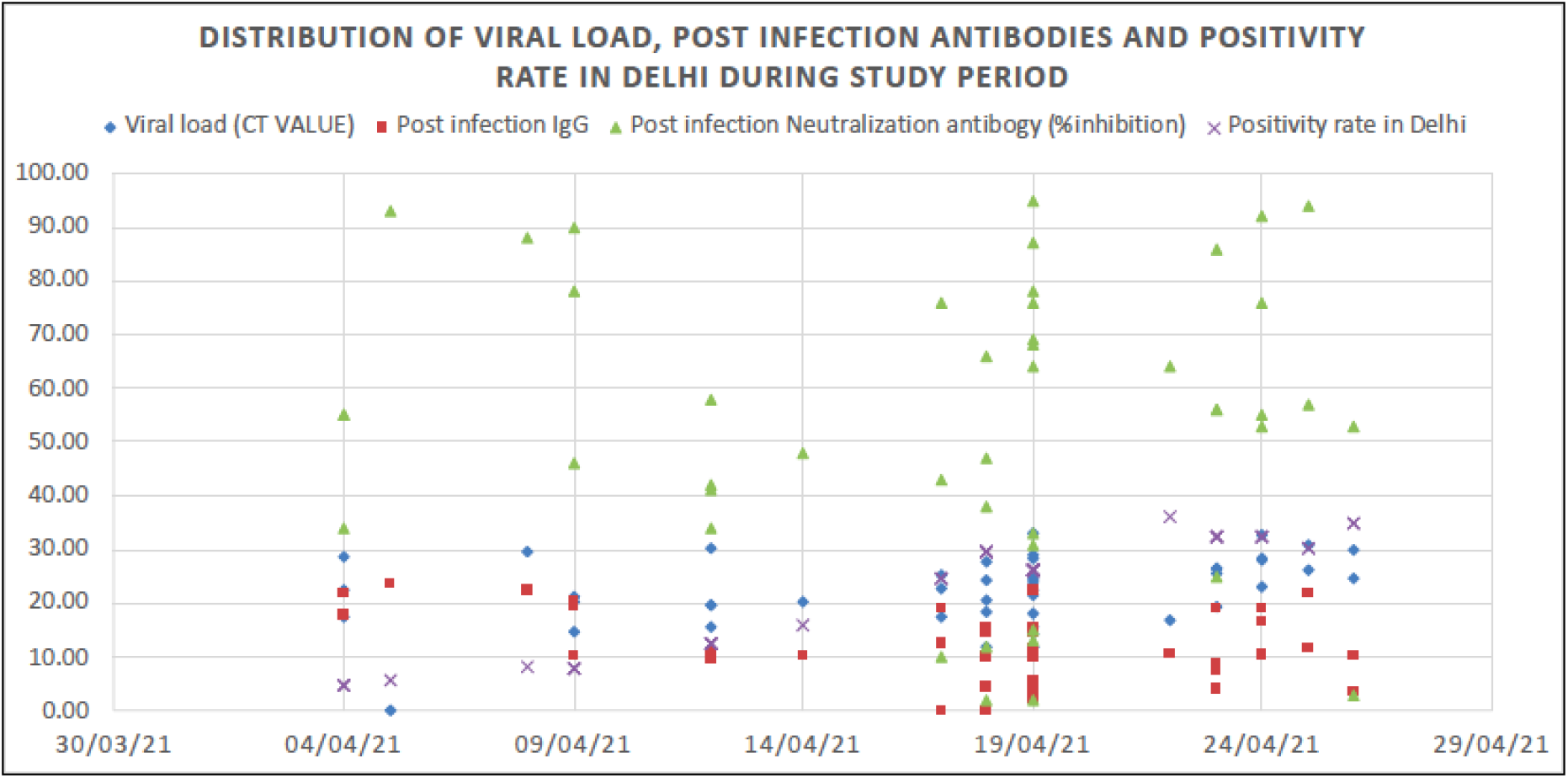
Distribution of study population, viral load (CT value), post infection IgG and neutralization antibody levels and the distribution of positivity rate in Delhi during the same time period.

## Discussion

Our prospective observational study documents the clinicogenomic analyses of largest number of vaccine breakthrough infections in HCWs. There was higher overall occurrence of infections, in the unvaccinated group (21.46%) as compared to the vaccinated subjects(9.52%), during the second wave in Delhi. In vaccinated group the infections were significantly higher in partially vaccinated (13.65%) than fully vaccinated group (8.62%) demonstrating the effectiveness of the double dose of vaccine. The study in healthcare workers at Christian Medical college, Vellore in India has reported BT infections in 9.6% fully vaccinated, 10.6% in partially vaccinated and 27.2% in unvaccinated HCWs.^15^ Their cohort included HCWs who received either ChAdOx1 nCoV-19/Covishield or BBV152/Covaxin. Other studies mostly report the BT infections after vaccination with BNT162b2 mRNA vaccine (Pfizer). Hall VJ et al, have reported 3.8% BT in HCWs vaccinated with BNT162b2 mRNA vaccine as compared to 38.5% in unvaccinated cohort from England.^16^ In another study by Keehner J from California have reported a low positivity rate of 0.05% after 2 doses of vaccination with BNT162b2 mRNA vaccine in HCWs.^17^ Benenson S et al., have reported BT in 6.9% of the vaccinated HCWs and 28.2% of the unvaccinated subjects.^18^ The data from three single-blind randomised controlled trials—one phase 1/2 study in the UK (COV001), one phase 2/3 study in the UK (COV002), and a phase 3 study in Brazil (COV003)—and one double-blind phase 1/2 study in South Africa (COV005) have shown that the overall vaccine efficacy 14 days after the second dose was 66·7%.^19^ The findings in our study shows similar efficacy 14 days after the second dose (60%).

There was predominance of BT among the doctors and nurses as compared to the technicians, non-medical staff and GDA. There was no difference in the infection rates in these categories in the non-vaccinated group. The predominance of BT in doctors and nurses indicates higher risk in the immediate care givers owing to the exposure to infected patients and close proximity during the patient care. There is no published data of BT infections in different categories of HCWs.

Higher severity of infection was also reported in the unvaccinated group. Clinically mild infection with fever and mild sore throat was the most common presentation in both the vaccinated and non-vaccinated groups with no severe disease or mortality. Similar clinical findings were also documented by Philomina *et al*., in breakthrough infections in 6 people receiving 2 doses of ChAdOx1 nCoV-19/Covishield vaccine in Kerela.^20^ In the recent study of breakthrough infections from Delhi, including a mixed cohort of HCWs, 10 received ChAdOx1 nCoV-19/Covishield vaccine and 53 received BBV152/Covaxin and none had severe infection.^21^ Thus, two doses of ChAdOx1 nCoV-19/Covishield vaccine offer protection against moderate to severe COVID-19 disease.

The serological analysis showed significantly higher antibody response in fully vaccinated HCWs as compared to partially vaccinated ones. Single dose recipients had higher BT infection rate and lower humoral response, predisposing them moderate to severe clinically morbidity. Earlier one case of breakthrough COVID-19 infection after full-dose administration of CoronaVac vaccine was reported from Indonesia by Zulvikar et al.^22^

There was predominance of B.1.617 lineage in a phylogenetic examination of 46 vaccinated COVID-19 positive samples collected in the months of April and May 2021 coinciding with the massive surge of cases in Delhi. The Delta variant was predominant in nearly 70% of the samples. As B.1.617.2 is dominating the numbers here, this suggests that there is a higher vaccine breakthrough risk with B.1.617.2 as compared to B.1.617.1 and B.1.1.7. Similar findings have recently been reported in another study.^21^ It is known that B.1.617.2 is characterised by 3 spike mutations L452R, T478K and P681R.^7^ B.1.617.1 is characterized L452R, E484Q and P681R.^23^ Similar predominance of B.1.617.2 in vaccinated individuals, was also reported from another tertiary care centre in Delhi during the surge of infections during March and April.^6^ The variants of concern B.1.617.1 partially impairs neutralizing antibodies elicited by BNT162b2 vaccine.^24^ Similar evasion for ChAdOx1 nCoV-19 (Covishield) vaccine induced antibodies is possible by the B.1.617.2. It confers evasion of elicited antibodies owing to the L452R mutation causing infections in individuals with pre-existing immunity from vaccines/natural infection. The RBD mutation T478K in the RBD is unique to B.1.617.2 facilitates antibody escape.^25^

### SARS-CoV-2 genetic variants associated with immune escape (Immune escape variants)

Despite the existence of several RBD mutations, studies to examine the capability of vaccinee sera to neutralise circulating SARS-CoV-2 variations showed that strains such as remain potently neutralised. Several RBD-specific antibodies can bind only the open spike protein^24^ and it has been discovered that D614G renders the spike protein more sensitive to neutralising antibodies by increasing the likelihood of the open conformation occurring.^25^ Nevertheless, other circulating SARS-COV-2 variations escape vaccine-induced humoral immunity.^26^ The L452R mutation was found in roughly 93 percent of the vaccinated COVID-19 positive samples, indicating that this mutation was positively selected. Leucine-452 is located in the RBD receptor-binding motif, at the point of direct interaction with the ACE2 receptor. Its substitution with arginine is expected to result in substantially greater receptor affinity as well as escape from neutralising antibodies.^27^ The structural investigation of RBD mutations L452R and E484Q, as well as P681R at the furin cleavage region, revealed the possibility of enhanced ACE2 binding and the rate of S1-S2 cleavage, resulting in improved transmissibility. The same two RBD mutations resulted in reduced binding to selected monoclonal antibodies (mAbs), decreasing mAb neutralising capacity, according to an Indian study.^28^ Considering the immune escape conferred by these RBD mutations, needs further survey.

Here we report the alarming rise in the SARS Cov2 variants causing breakthrough infections in vaccinated healthcare workers. Further immune surveillance and characterization of the variants is warranted to understand the pathophysiology and design measures to curb the spread of the variants.

## Data Availability

the data relevant to the study are available

